# The effective reproductive number (Rt) of COVID-19 and its relationship with social distancing

**DOI:** 10.1101/2020.07.28.20163493

**Authors:** José Alexandre Felizola Diniz-Filho, Lucas Jardim, Cristiana M. Toscano, Thiago Fernando Rangel

**Affiliations:** Departamento de Ecologia, Instituto de Ciências Biológicas, Universidade Federal de Goiás (UFG), Brasil; Instituto Nacional de Ciência & Tecnologia em Ecologia, Evolução e Conservação da Biodiversidade, UFG, Programa DTI/CNPq, Brasil; Departamento de Saúde Coletiva, Instituto de Patologia Tropical e Saúde Pública (IPTSP), UFG, Brasil

**Keywords:** Social isolation, time series analysis, effective R, COVID-19, real-time intervenion

## Abstract

The expansion of the new coronavirus disease (COVID-19) triggered a renewed interest in epidemiological models and on how parameters can be estimated from observed data. Here we investigated the relationship between average number of transmissions though time, the reproductive number *Rt*, and social distancing index as reported by mobile phone data service *inloco*, for Goiás State, Brazil, between March and June 2020. We calculated *Rt* values using *EpiEstim* package in R-plataform for confirmed cases incidence curve. We found a correlation equal to -0.72 between *Rt* values and isolation index at a time lag of 8 days. This correlation is also significant for half of the cities of the State with more than 90,000 people, including the 3 largest ones (and the 7 cities with significant correlations account for 43% of the population of the State). As the *Rt* values were paired with center of the moving window of 7 days, the delay matches the mean incubation period of the virus. Our findings reinforce that isolation index can be an effective surrogate for modeling and epidemiological analyses and, more importantly, helpful for anticipating the need for early interventions, a critical issue in public health.

## 1. Introduction

The global expansion of the new coronavirus disease (COVID-19) triggered a great and renewed interest in epidemiological models to understand temporal and geographical patterns of expansion of the pandemics and to use such models to guide real-time decision making to mitigate its spread (Adam, 2020; Jewell et al. 2020). As so, it is of utmost importance to better understand clinic, immunologic and epidemiological characteristics of the new coronavirus (SARS-CoV-2) that causes COVID-19 and how these epidemiological features can be translated into statistical parameters estimated from real-time observed data to provide accurate predictive models. In the core of such discussions lies the basic reproduction number (R_0_), which reflects the average number of new infections generated from an initial infection in a susceptible population (Ridenhour et al., 2014). This parameter synthesizes, under a more realistic view, incubation and transmission periods of the infectious agent (Lauer, 2020; He et al. 2020), but actually it is not constant though time. During an epidemic, changes in the effective or realized infection transmission through time (here denoted as *Rt*) also reflect the demographics of the host-pathogen interaction, which is regulated by several extrinsic environmental effects (mainly population density and social contact) (Badr et al., 2020; Flaxman et al., 2020). Thus, in the particular case of COVID-19 pandemic, for which no specific therapeutic or preventive strategy is yet available, the downward shifts in *Rt* is the expected outcome of non-pharmaceutical interventions, including social distancing measures, sanitary measures and use of masks, to reduce the number of transmissions. These interventions flatten the epidemic curve, allowing healthcare systems to prepare and manage the expected cases of disease, while waiting for a vaccine development, its high-scale production and availability to wide population.

Although it is relatively simple to estimate the infection *Rt* based on incidence data and estimates of serial interval (Abbott et al., 2020), it is much more difficult to explain its continuous temporal variation given the implementation of interventions, in particular social distancing measures (Badr et al., 2020). Given the characteristics of COVID-19 such as its large proportion of oligo and asymptomatic cases, the ability to be transmitted by asymptomatic individuals, and its rapid spread, it is impossible to accurately identify the number of infected individuals early in the epidemic. As such, it is important to have surrogates for real-time tracking of the *Rt* dynamics. The broad-scale real-time monitoring of mobility derived from mobile phones has been reported to significantly correlate with decrease in the number of COVID-19 cases and increased social distance in the population (Badr et al., 2020). Particularly in extremes of the epidemic, social distancing estimated by mobility indicators have been one surrogate of the *Rt* dynamics (Flaxman et al., 2020; Kupferschmidt, 2020; Mellan et al., 2020). However, in addition to these extremes, it is interesting to assess whether there is a more continuous relationship between mobility indicators and *Rt* over time, as this would allow closer and timely monitoring of disease transmission, which may guide public health responses tailored at the local level (Badr et al., 2020).

Here we investigate the relation between *Rt*, estimated from incidence curves of confirmed cases, and social distancing index from mobile phone data in Goiás State in Central Brazil and its major cities. The first COVID-19 cases were identified in Goiás in mid-March and, shortly after, while there were only few imported confirmed cases notified to the local surveillance system, the State Government implemented strict social distancing measures, including cancellation of events, school and workplace closures, closure of commercial establishments and services, except for essential services (Silva et al., 2020) (see also https://medidas-covidbr-iptsp.shinyapps.io/painel/). After these initial measurements, social distancing was relaxed and isolation indicators gradually decreased, thus providing an opportunity to evaluate the continuous relationship between *Rt* and social distancing index.

## 2. Methods

Goiás state has approximately 7 million people living in its 246 municipalities (SEGPLAN, 2020). We considered epidemiological and mobility data from all municipalities in the state. Isolation indicator is given as 1 - MI, where MI an index of social mobility estimated as the percentage of electronic devices (eg., mobile phone, tablets) leaving a radius of 200 meters from a location considered “home” by *InLoco* app users (https://mapabrasileirodacovid.inloco.com.br/pt/). Isolation indicador was obtained for each municipality in Goiás state and overall isolation was obtained by population-weighted average. Epidemiological data for COVID-19, including confirmed cases and deaths, were obtained from the Goiás State Health Department (SES-GO, available at http://covid19.saude.go.gov.br/). The study period ranged from March 16 to June 6, 2020. We estimated the *Rt* values based on the incidence curve of confirmed COVID-19 symptomatic cases reported in the study period representing a total of 14,751 cases.

We used the package *EpiEstim* of the R language (R Core Team, 2020; Cori et al., 2013) to analyze incidence by dates of symptoms. Despite the significant delay for case confirmation in the system, of approximately 7-10 days in Goiás state, we considered cases by date of symptom onset, therefore, we have enough replications to represent the changes disease incidence. We modeled *Rt* through time as a Poisson distribution whose the expected amount of new infected individuals is, on average, 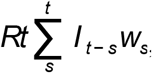, where *w*_*s*_ is the probability distribution of time until secondary infection (Cori et al., 2013), approximated by a discretized Gamma distribution (µ = 5.2 and σ = 0.6 for *w*_*s*_) (Abbott, et al., 2020; Kupferschmidt, 2020; Ganyani et al., 2020). The I_t-s_ is the total incidence up to time *t*. We estimated the posterior distribution of *Rt* by assuming a prior Gamma distribution (a = 1, b = 5). The values of *Rt* were calculated using a moving average of 7-day window.

Although more sophisticated methods are available to account for delays of disease notifications and eventually sub-notifications in the estimation of real-time *Rt* (Gostic et al., 2016), we simply truncated the end of the distribution of cases as our goal is not to have a real-time estimate of *Rt*, but rather to evaluate the effectiveness of isolation indicator as a proxy of *Rt* shifts throughout the time series.

The isolation indicator for the entire state were correlated to the *Rt* values, initially, using standard Pearson correlations. We paired the *Rt* values to isolation indicator assuming that each *Rt* reflects the centre of each time window. However, considering date of symptom onset and the fact that infection transmission take place in average 5 days earlier (Lauer et al., 2020; He et al., 2020), we also calculated these correlations considering different time lags to evaluate its behavior (from 1 to 12 days). Because of the strong temporal autocorrelation in both time series, especially in the *Rt* values (as they are calculated using overlapping moving windows), it is not appropriate to apply a standard statistical test due to inflated Type I error. We then tested the correlation between *Rt* and isolation indicator using the Dutilleul’s method to calculate the effective degree of freedom based on temporal Moran’s *I* correlograms taking into account positive autocorrelation in both time series (Dutilleul et al., 1993; Legendre and Legendre, 2013).

We repeated the above calculations of *Rt* and its correlation with isolation index for each of the 246 cities of the State. We also evaluated how correlation is affected by uncertainty in serial interval of COVID-19 by repeating it 1000 times randomly varying this value between 3.5 and 6.5 (see Nishiura et al. 2020).

## 3. Results

The early intervention by State Government in March declaring a quarantine which resulted in a significant increase in the isolation indicator from about 25% to ca. 55% within a one-week period (Fig. 1A). However, in March there was a very small number of confirmed cases, so as expected the COVID-19 epidemic curve grew very slowly in the state (as well as in surroundings states encompassing the entire Central region of Brazil) from mid-March to mid-May 2020. After this 2 month-period, isolation indicator decreased steadily together with the observed increase of case counts and disease transmission from mid-May onward.

**Figure 1.**
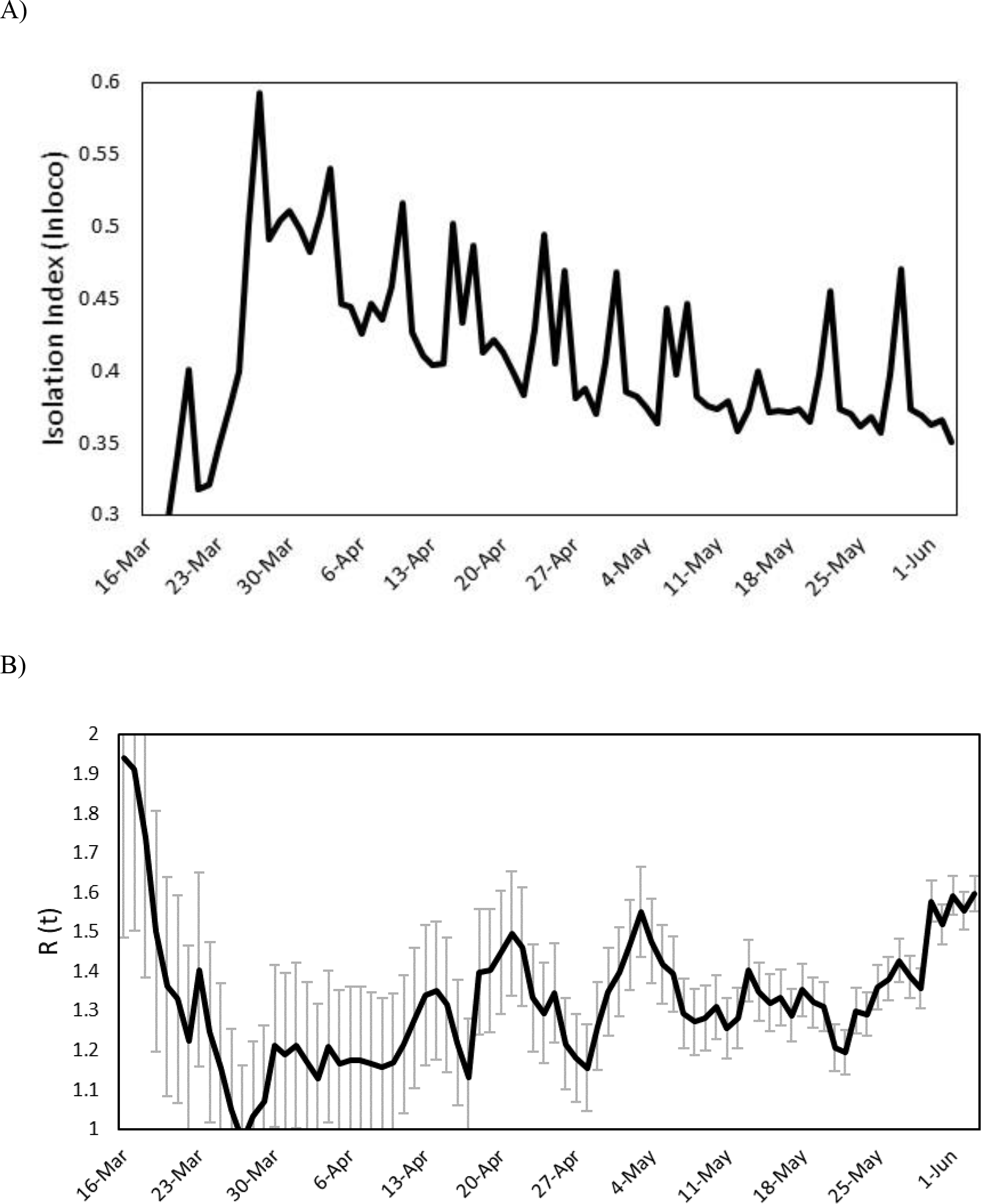
Times series of Isolation index (*InLoco)* (A) and mean reproduction number (*Rt*) though time in Goiás State (B) between March – June 2020, with respective 95% confidence intervals.

The time-series of *Rt* (Fig. 1B) shows the effects of the early and strict social distancing policies in Goiás State in mid-March 2020, reducing the *Rt* from around 2.0 to about 1.0 in middle April 2020. However, it is possible to observe that *Rt* increases slowly from this point on, reaching 1.4 - 1.5 in late May/early June 2020.

However, even though overall pattern of the time-series are qualitatively similar, when we correlate *Rt* and the isolation indicator at a fixed data a relatively low correlation (r = -0.281) is observed. However, it is indeed necessary to consider a time-lag to reveal a relatively high negative correlation, and it is possible to find a much higher correlation of -0.636 at time lag 8. Hence, the number of transmissions is associated with isolation indicator of 8 days before, as we used a 7-day time window, it closely matches the 5 average days of incubation of the virus. The relationship may be slightly non-linear (Fig. 2), although Spearman rank correlation is not that different from Pearson’s correlation (−0.645). A log-transformation in both variables increased the correlation to -0.685.

**Figure 2.**
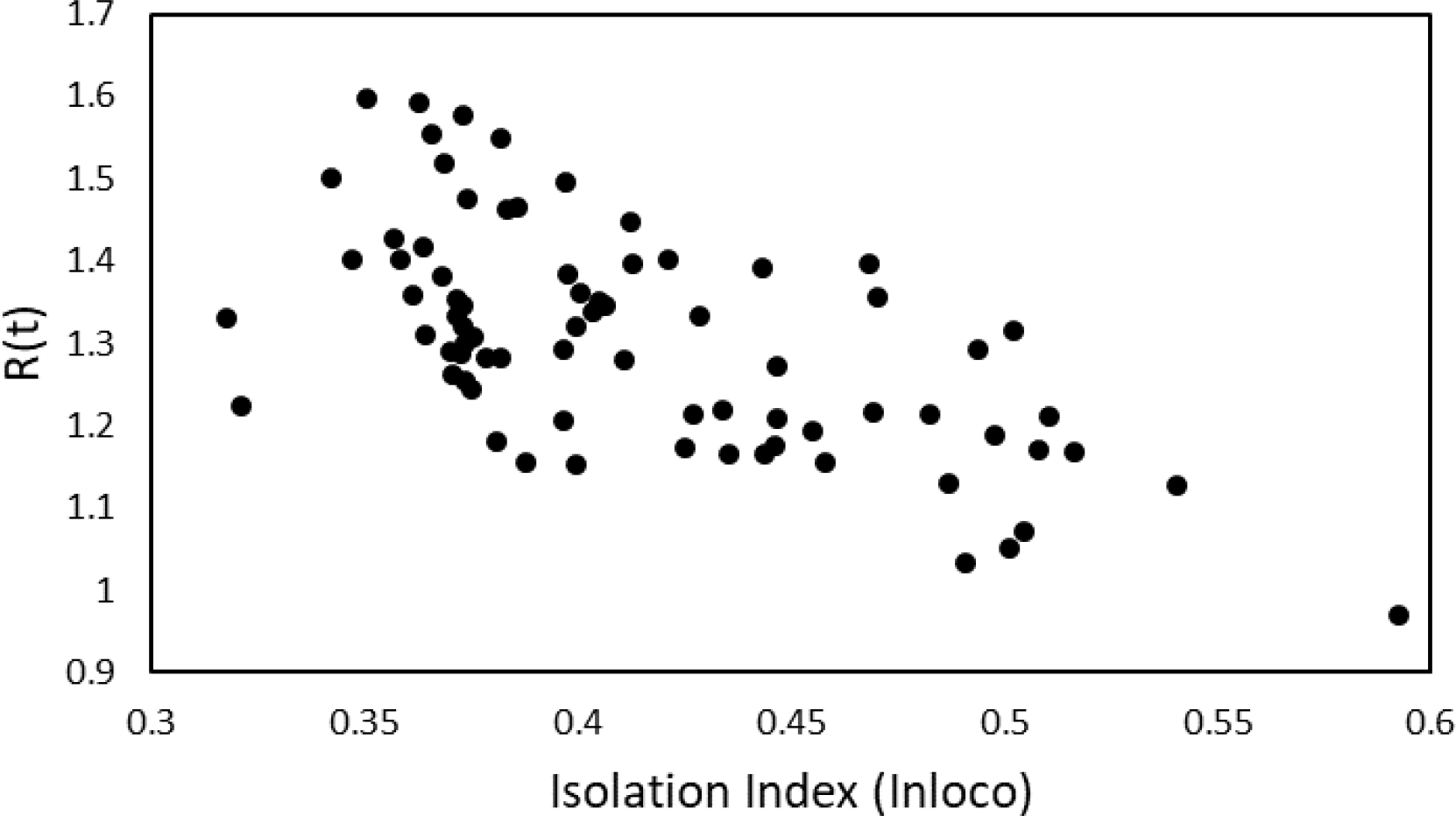
Negative relationship between Reproduction number *Rt* and isolation index measured by *InLoco*, Goiás State, between March and late May. The correlation is equal to 0.696 at original scale and, due to a slightly non-linearity, it increases to 0.721 after log-transforming both axis).

The first class autocorrelation using Moran’s I autocorrelation coefficient in both variables is around 0.5, decreasing to zero after ca. 21 days, and thus Dutileull’s method suggest that correlation between the two series should be effectively tested with about 18-20 degrees of freedom (and even in this case these correlations are significant at P < 0.01). For the analyses of the cities, these degrees of freedom vary a bit as a function of the autocorrelation in each series, but in general values of correlation lower than -0.3 tend to be statistically significant at P < 0.05.

Although there is a non-linear relationship between correlation between *Rt* and isolation index and the serial interval used (see Nishiura et al. 2020), even lower values of serial interval around 3.5 days results in a significant correlation of -0.60 for the entire State, which does not qualitatively affect our conclusions.

Significant correlations between *Rt* and isolation index were found for only 7 out of the 246 cities in the State. Actually, the overall pattern for the State can be attributed mainly to the three cities with more than 250,000 people, Goiânia (the capital), Aparecida de Goiânia and Anapolis (Fig. 3). Out of the 15 cities with more than 90,000 people, correlation was observed for 7 of them, mostly situated in the central part of the State. These 7 cities with significant correlations encompass ca. 43% of the State population.

**Figure 2.**
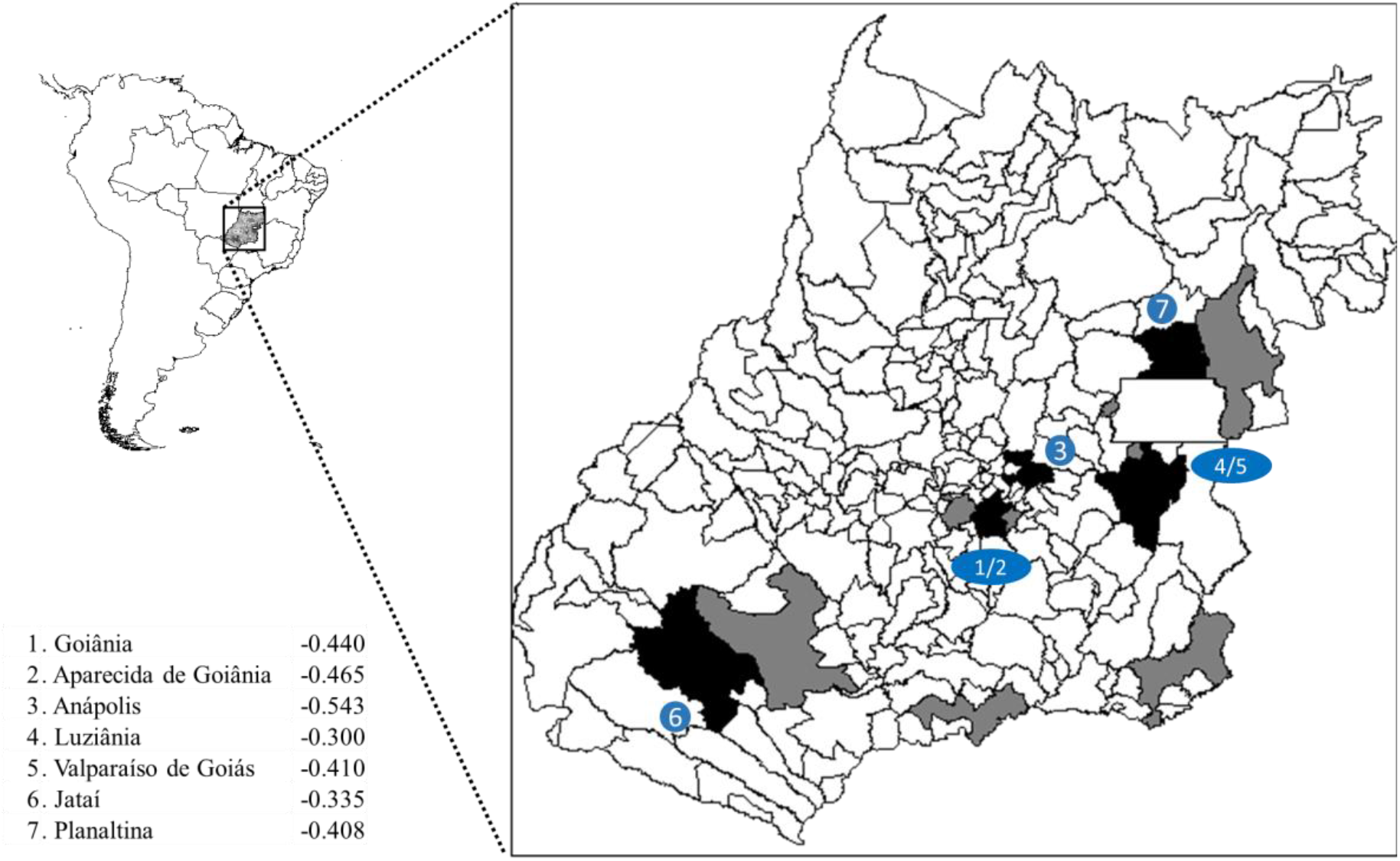
Goiás State in Central Brasil, showing the main municipalities (cities are capitals) with more than 90,000 people (gray areas) and those with significant correlations between isolation index and *Rt*. The numbers refers to each cities, with the estimated correlations shown in the table at lower left.

## 4. Discussion

There is a real interest to evaluate the local dynamics of disease transmission, as this is the main parameter that drives the spread of COVID-19 in a region. Several analyses have shown, as expected, that social distancing policies effectively reduce the reproduction number (*Rt*) of disease spreading (Badr et al., 2020; Flaxman et al., 2020). However, it is actually difficult to evaluate the true effects of such measures in a more realistic fashion, as there may be several confounding and interactions between broad-scale isolation related to mobility and more local and “behavioral” aspects at very local and individual scale, including hygiene measures, mask use and social awareness as a result of the pandemic (Chu et al., 2020; Kucharski et al., 2020), as well as a more effective contact tracing and early detection of cases (Peto et al., 2020).

We identified for Goiás State a relatively high negative correlation of -0.685 at log-scale) between the estimated *Rt* and isolation indicator obtained from mobile phone data. Correlation for *Rt* based on death incidence curve are slightly slower, for a larger time lag, which is expected because of the relatively smaller number of cases, as well as due to a more complex series of events from transmission to death (including a strong effect of age-structure in hospitalization in intensive care facilities). For transmissions, anyway, our empirical results match those reported for São Paulo State, where the *Rt* decreases from about 2 to 1 after strict social distancing interventions were implemented in mid-March 2020. Similar findings were also reported for broad-scale analysis of other countries (Badr et al., 2020; Kupferschmidt, 2020). However, our results show that this high correlation between *Rt* and isolation indicator suggests a more continuous relationship between these variables, as recently calculated for United States (Badr et al., 2020), rather than a more discrete variation of extremes (i.e, if isolation increases to 50% the *Rt* decreases to 1.0, as shown for São Paulo state (Ganem et al., 2020), France (Roques et al., 2020), and Europe in general (Flaxman et al., 2020), and used for initial modeling in Brazil (Mellan et al., 2020).

Even more interestingly, the negative correlation between isolation and *Rt* appears only if a time-lag is created, which is indeed expected considering current knowledge of COVID-19 disease and in particularly its incubation time. This correlation may seem clear to our case in Goiás, perhaps due to the variability of time series, with an initial increase in the isolation indicators shortly after the start disease transmission locally, followed by a more continuous reduction in the isolation indicator, and finally by a stabilization at around 38% in late May. Even so, it is also important to note that this correlation between *Rt* and isolation indicator must be better detected in early phases of the COVID-19 expansion, and it will tend to disappear as the epidemic size increases and the peak of transmissions occurs because Rt will be also more strongly affected by the reduction of number of susceptible individuals in the population.

Another important issue to be discussed is the spatial patterns in the correlation between *Rt* and isolation, which seems to be associated to population size in cities, as expected by its association with increase in number of cases (i.e., Coelho et al. 2020; Berg et al. 2020). Out of the 15 cities, correlations between Rt and isolation index was observed for 7 of them, all with almost 100,000 people. Of the four largest cities in the State, only for the fourth one (Rio Verde, with about 230,000 people) there is no correlation between Rt and isolation index. Actually, this is an interesting case as this city was one of the first cities in Goiás to confirm a case (just after the capital) so we are aware that strong control measures were declared since the beginning of the pandemics. Now the cities is actually the third one in the State in terms of number of death and a huge number of confirmed cases, but this was due to a late superspreading event in large meat processing plants, in late June. It is interesting to note that a significant correlation is observed in the neighbor city, Jatai.

The other important spatial component of diffusion in Goias state is related to the proximity with the Federal District (DF), which encompass the capital of the country, Brasilia (the retancgle inside the central-eastern portion of the State), with almost 4 million people. Thus, although political and administration measure in respect to COVID-19 were independent of in DF, it is not possible to neglect its effect of the neighbouring regions, which also concentrates another large part of the population in Goias state. Since the beginning of the pandemics, these cities surrounding the DF tended to adopt epidemiological control of borders that reduced their effect and slowed down the growing of the infection, especially the largest ones (i.e., Formosa, on the eastern side of DF in Fig. 3). Even so, again half the cities with more than 100,000 people surrounding the DF showed significant correlations between *Rt* and isolation index.

So, the cases of Rio Verde and Formosa seems to be informative in the sense that strong local controls, at least in early phases of the pandemics, tend to buffer the broad-scale correlation between population mobility and number of transmission and further events. Although epidemics started in all these cities early, in some of them contact tracing and border control seems to be more effective to minimize early growth of number of cases.

In conclusion, about 50% of the variation in the mean number of transmissions expressed by *Rt* in the Goiás state is explained by broad-scale isolation indicator measured by mobile phones. Although there is a relatively large amount of unexplained variation, we do not have additional variables related to more local measures that could improve our understanding of disease transmission. Considering the amount of variation explained and the randomness of the residuals of the relationship between isolation and *Rt*, we understand that isolation indicators obtained from mobile phones can be a good surrogate for COVID-19 transmission in a given population, and can be used for modeling purposes as well as routine epidemiological analyses. More importantly, it and can be quite useful for real-time public health policies, anticipating the need for early interventions, an important and critical issues in public health.

## Data Availability

.

## Acknowledgments

Authors thank National Council for Scientific and Technological Development (CNPq) for productivity fellowships and grants. Our work on simulation and population modeling has been continuously supported by National Institute of Science and Technology (INCT) in Ecology, Evolution and Biodiversity Conservation (J. A. F. D.-F. and T.F.R), supported by CNPq and FAPEG. CMT is a researcher of the Institutes for Health Technology Assessment (IATS), also supported as a INCT by CNPq. We thank Goias State and Goiania city Health Departments for support and access to original data.

## Author’s contributions

J.A.F.D.-F., C. M.T. and T.F.R designed the work and J. A. F. D.-F. did the preliminary analyses; LJ implemented the code for *EpiEstim* and did the first analyses, whereas T. F. R. processed all data on social index; all authors contributed to the writing and reviewing of the final submitted version.

